# Bump2Baby & Me+ (B2B&Me+): Protocol for a multi-country, European implementation project to reduce the incidence of gestational diabetes mellitus and improve maternal and child health

**DOI:** 10.64898/2026.07.09.26357618

**Authors:** Sharleen L O’Reilly, Thérèse McDonnell, Linda Reme Sagedal, Mercedes G Bermúdez, Florian Herrmann, Hanna Jasiak-Jóźwik, Nina Cecilie Øverby, Hilde Kristin Refvik Riise, Fionnuala McAuliffe, Helle T Maindal, Laura Headley, Cristina Campoy, Sebastian Kwiatkowski, Ragnhild B. Strandberg, Anju Rawal, Katie Angotti, Hannah Foley, Lucy Murphy, Lin Chen, Quentin Le Cornu, Marjolein M Iversen, Timothy Skinner

## Abstract

**Background:** Gestational diabetes mellitus (GDM) affects 1-in-7 pregnancies globally and is associated with significant short- and long-term health consequences. Although health behaviour change interventions can effectively reduce these risks, a significant implementation gap exists in translating this evidence into routine practice. Bump2Baby and Me (B2B&Me) was a mobile health (mHealth) coaching intervention provided to women at-risk of GDM from early pregnancy through to 1-year postpartum. B2B&Me Plus (B2B&Me+) aims to refine, implement and evaluate the implementation of this personalised intervention across 4 countries (Ireland, Spain, Poland and Norway) with differing health systems and population profiles.

**Methods:** This study employs a hybrid type 3 implementation-effectiveness design using a non-randomised ABA block approach within a longitudinal cohort. Participants will be screened using the Monash machine learning GDM screening tool (MMLGDST). During the intervention (Block B), women at risk of developing GDM will be offered access to a smartphone-based coaching application featuring 1:1 synchronous sessions, asynchronous text and video messaging along with a Bluetooth-enabled weighing scale for self-monitoring. Support continues from early pregnancy through to nine months postpartum.

The study’s primary objective is to evaluate reach, adoption, implementation and maintenance of the B2B&Me+ intervention programme when delivered within routine maternity care. Implementation success will be assessed using the RE-AIM framework, while secondary outcomes will assess intervention effectiveness. The study will examine population-level uptake at each site, evaluate the benefits and costs of implementation across varying contexts, and analyse how four different referral methods, randomised at the site level, affect uptake. In addition, a European implementation toolkit will be developed to provide health services with scalable strategies to bridge the evidence-to-practice gap.

**Discussion:** This study will contribute to a growing literature on the implementation of a successfully trialled mHealth intervention in a real-world context. Understanding variation in both intervention and implementation success within a routine maternity care context across diverse settings will inform the development of an implementation toolkit to support health services in reducing the incidence of GDM and improving maternal and child health outcomes.

This study was registered with ClinicalTrials.gov (NCT07189221) on 28 August 2025 (https://clinicaltrials.gov/study/NCT07189221?cond=diabetes)

**Contributions to the literature:**

- B2B&Me+ is the first multi-site European study testing how a mobile health coaching programme for women at-risk of gestational diabetes can be delivered within routine maternity care across four countries.
- The study tests four strategies for inviting women to join the programme, with the intent to identify which strategy most effectively reaches and engages them in practice.
- Using established implementation science frameworks, B2B&Me+ examines what drives or hinders delivery across four diverse European health systems, generating evidence applicable beyond the study settings.
- Findings will directly inform a practical European toolkit to support maternity services in adopting the programme at scale.

## BACKGROUND

Gestational diabetes mellitus (GDM) affects approximately 1-in-7 pregnancies worldwide and is associated with significant short and long-term health consequences for both mother and baby (1, 2). Although GDM typically resolves after delivery, it remains a strong predictor of type 2 diabetes (T2D), with women who have had GDM facing a 10-fold increased risk of developing it much earlier than the average person (3). Rates of GDM are rising, with this increase attributed to factors such as higher maternal age, excess weight and health behaviour change factors such as poor diet, low physical activity, sedentary behaviours, and stress (2, 4). Higher body mass index (BMI) is the most significant modifiable risk factor for GDM (5). In addition, excessive gestational weight gain (GWG) is associated with a range of adverse maternal and neonatal outcomes (6–10). Considering the consequences of GDM and excessive GWG, it is important to mitigate risk and improve maternal and neonatal outcomes.

Diet and physical activity interventions during pregnancy can effectively reduce GWG, GDM risk, and other pregnancy complications (11–13). Systematic review and meta-analysis concluded that interventions using diet, physical exercise, metformin,, and/or myoinositol compared to usual care reduced the incidence of GDM (14). Despite greater awareness of health behaviour change’s importance, it remains challenging to maintain engagement within traditional behaviour modification programmes. Mobile health (mHealth) interventions offer a promising solution by providing scalable, cost-effective, and accessible support for health behaviour change modification. The Horizon 2020-funded Bump2Baby and Me study on 865 women at risk of developing GDM demonstrated promising results (15). The intervention group had a lower incidence of GDM (14.0% vs. 20.0%, P=0.027), had lower 12-month fasting glucose (5.01 vs. 5.17 mmol/L, P=0.009), as well as lower insulin (102.09 vs. 141.17 pmol/L, P=0.034) and triglycerides (1.69 vs. 1.81 mmol/L, P=0.038), and higher HDL cholesterol (1.48 vs. 1.43 mmol/L, P=0.007). Any breastfeeding at 6 weeks postpartum was higher in the intervention group (88.1% vs 81.2%, P=0.02), as was exclusive breastfeeding at 3 months (60.6% vs 51.9%, P=0.047). Self-weighing during either pregnancy or postpartum was associated with a reduction in BMI of 2.34 kg/m² (-3.68 to -0.99) and during both periods with a reduction of 3.45 kg/m² (-4.68 to -2.21) (15). Limited evidence exists on translating this evidence into routine antenatal and postpartum care.

Guidance on delivering mHealth coaching programs or digital health interventions covering health outcomes, cost-effectiveness, and long-term sustainability are lacking (16). Health services face a daunting task when implementing such interventions with limited resources. Staff shortages and high GDM rates compound these issues, resulting in women not receiving optimal, guideline-led care when they need it most. Therefore, greater emphasis must be placed on implementation research to ensure more coherent global evidence and the alignment of such interventions within the context of underlying system constraints (16).

A structured approach to evaluating how interventions perform when translated into routine practice rather than under controlled trial conditions is critical for research implementation. One such established framework is RE-AIM (Reach, Effectiveness, Adoption, Implementation, Maintenance) (17), which has been extensively applied across public health and health services research to evaluate intervention translation. While the areas of reach and effectiveness are commonly reported in detail, adoption, implementation and maintenance remain under-reported (18). The B2B&Me+ project aims to address this gap by testing the real-world effectiveness of the mHealth coaching programme within regular maternity services across four European countries (17, 18). Implementation in four different locations that vary greatly in terms of health systems, including national GDM screening pathways and referral processes, and demographic profile such as ethnicity and socioeconomic status, will improve the understanding and delivery of implementation. This geographical diversity will provide valuable insight on which factors are common barriers, and which are likely to be country specific. B2B&Me demonstrated good penetration and uptake within the context of a clinical trial. However, clinical trials are not designed to capture the broader service-level impact, the real-world strain on hospital resources, staff workload, and operational workflows. Therefore, to assess the challenges, advantages, and impacts associated with integrating B2B&Me+ into routine healthcare services, this study will examine population-level uptake at each location, evaluate the benefits and costs of implementation across varying contexts, and analyse how different referral methods affect uptake.

### Conceptual Framework and Rationale

The Exploration, Preparation, Implementation, Sustainment (EPIS) framework (19, 20) is B2B&Me+’s overarching conceptual framework, guiding the study design and structuring our understanding of the multi-level factors that influence implementation across four diverse health systems. EPIS describes four sequential phases of evidence-based practice implementation and maps these against both the outer context (national health systems, policy environments, population-level GDM burden, and digital health infrastructure across Ireland, Spain, Poland and Norway) and the inner context (each maternity site’s organisational readiness, staff capacity, existing antenatal care pathways, and attitudes towards mHealth tools). Bridging factors, particularly the consortium partnership structure and the involvement of Liva Healthcare A/S as the mHealth platform provider, connect these contexts and support consistent delivery across sites. The B2B&Me+ mHealth coaching programme constitutes the innovation factor, characterised by the evidence base from the original B2B&Me RCT, the Monash gestational diabetes screening tool, and the personalised health coaching platform.

The EPIS phases map directly onto B2B&Me+ study activities. *Exploration* encompasses the original RCT evidence, the implementation gap in translating mHealth behaviour change interventions into maternity care, and this protocol development. *Preparation* is operationalised through pre-implementation contextual mapping at each site, characterising inner and outer context barriers and facilitators prior to study commencement, and informing site-level preparation including staff training and mHealth app adaptations. *Implementation* encompasses the study itself, a non-randomised ABA block design, where women are screened using the MMLGDST and invited to the B2B&Me+ mHealth coaching programme using one of four randomised referral strategies within the intervention period (Block B). This block is then compared with two usual care blocks before and after. The rationale for comparing referral approaches (point-of-care referral and information leaflet, both with and without active follow-up) was informed by prior evidence that active follow-up strategies yield higher proportional enrolment while more passive strategies achieve greater absolute penetration (21). *Sustainment* is addressed through the co-design of a European implementation toolkit, which will translate study learnings into scalable guidance for health services considering future adoption of the programme.

Normalisation Process Theory (NPT) (22) complements EPIS by providing the study’s qualitative analytical framework. Interviews and focus groups with health coaches, site coordinators, and clinical leads will be structured around NPT’s four constructs (coherence, cognitive participation, collective action, and reflexive monitoring) to ask what work is needed and by whom to make this innovation become routine, embedded and sustained within everyday maternity services. Findings from this analysis will directly inform the implementation toolkit, supporting understanding of the conditions under which B2B&Me+ might be normalised into routine maternity care in the future.

### Study Aims

Aim 1 (Primary — Referral Methods): To compare participation rates achieved by point-of-care versus information leaflet referral strategies among MMLGDST-identified at-risk women, hypothesising that point-of-care approaches will yield higher proportional participation (23).

Aim 2 (Primary — Implementation): To evaluate the reach, adoption, implementation and maintenance of B2B&Me+ across four European maternity services using the RE-AIM framework, with EPIS guiding exploration of the inner and outer context factors that explain variation across sites.

Aim 3 (Secondary — Effectiveness): To assess the effectiveness of B2B&Me+ on maternal and neonatal outcomes, hypothesising lower GDM incidence, reduced gestational weight gain, and improved breastfeeding outcomes in the intervention block compared with usual care, consistent with the B2B&Me RCT (15).

Aim 4 (Exploratory — Implementation Experience): To explore the implementation experience of health coaches, site coordinators and clinical leads using NPT as an analytical lens, generating evidence to inform the co-design of a European implementation toolkit.

## MATERIALS & METHODS

### Project Design

This protocol is reported in accordance with the Standards for Reporting Implementation Studies (StaRI) (17). (Supplementary file 1). This is a hybrid type 3 implementation-effectiveness study using a non-randomised, ABA intervention design nested within a longitudinal cohort. As a type 3 hybrid study, focus is primarily on implementation outcomes, with secondary outcomes assessing intervention effectiveness (24).

### Implementation Strategy

The study will compare the delivery of the MMLGDST and mHealth coaching referral during a 3-month intervention block (B) with two 3-month blocks of usual care, before and after the intervention (A blocks) (25). Recruitment to the study commenced on 1^st^ December 2025 and is scheduled to conclude on 31^st^ August 2026. Data collection is scheduled to be completed in January 2028, and results are expected to be reported by May 2028.

All participants will complete a baseline questionnaire and provide written informed consent to access medical records. While Block A participants will not undergo risk screening, their GDM risk status will be determined from the data provided. Within Block B, women attending the antenatal clinics across the four sites will be recruited either in-person or by phone. During the intervention block (B), women will provide written informed consent to be screened using the MMLGDST. Those screening positive to being at-risk of GDM will be provided with information about the B2B&Me+ programme and invited to provide additional written informed consent for the intervention.

Pregnant women attending their first/early antenatal appointments at participating sites will be assigned to a block based on the date of their visit. Gestation must be less than 24 weeks at enrolment; participants must be aged 18 years or older and able to provide informed consent. Women with a multiple pregnancy (twins, triplets, etc.), existing diabetes (Type 1 or Type 2) or early GDM are excluded. To be considered eligible for the intervention (Block B only), women must be identified as at-risk for GDM using the MMLGDST and must also own a smartphone. Additional exclusion criteria apply for the intervention group:

- Participation in another health behaviour change intervention study during pregnancy
- Inability to understand the language of the intervention (English in Ireland, Spanish in Spain, Norwegian in Norway, Polish in Poland)
- Severe mental illness, drug or alcohol abuse that would impair ability to participate
- Cancer (not in remission)
- Myocardial infarction in the last three months
- Not owning a smartphone capable of hosting the intervention app.

### Context

The assessment of context is important for implementation evaluation. The study will be conducted in four maternity hospitals across Europe:

1. National Maternity Hospital, Dublin, Ireland
2. University Hospital San Cecilio, Granada, Spain
3. Sørlandet Hospital Trust, Kristiansand, Norway
4. Pomeranian Medical University Hospital, Szczecin, Poland

These sites were selected to represent diverse healthcare systems, populations, and models of maternity care across Europe. Contextual barriers and facilitators that are linked to different roles, tasks, education, and skills of health and care professionals will be considered within the project and have their own dedicated work package. At the outset of the project, researchers will assess knowledge, attitudes and barriers that exist within the four study hospital sites to adopting the intervention and develop a tool to speed up this implementation process into the future. Staff within each hospital will be surveyed and interviewed to see what their awareness level is of related services that are available and the work associated with supporting women at risk of developing gestational diabetes. On completion of the intervention, a subset of intervention participants from each site will be invited to participate in an interview to explore their intervention experience as well as implementation barriers and enablers. Researchers involved in the recruitment and management of the study and mHealth coaches will also be invited to take part in an interview/focus group for the implementation analysis of the project.

### Intervention Referral

Within Block B, different referral methods will be tested every 2 weeks for women assessed as at-risk of GDM and meeting the eligibility criteria:

i. Point-of-care (POC) active referral: programme offered either in-person or by phone if the woman agrees to a phone consultation after the initial antenatal appointment. The participant is supported to start the sign-up process for the B2B&Me+ mHealth coaching app and the VitaDock+ Bluetooth scales app during this contact time.
ii. POC with follow-up phone call: As above, plus follow-up call within 48 hours to support sign-up.
iii. Leaflet provided: programme offered either in-person or by phone if the woman agrees to a phone consultation after the initial antenatal appointment. On enrolment, the participant is provided with an information leaflet about the sign-up process for both apps.
iv. Leaflet with follow-up call: As above, plus follow-up call within 48 hours to support sign-up.

The referral allocation will be site-stratified and randomised. The randomisation sequence will be generated by the project manager who is not involved in participant recruitment using computer-generated random numbers. Site coordinators will enrol participants to the appropriate intervention pathway based on the randomisation schedule. The sequence will be stored securely by the project manager and revealed to each site no more than two working days before the commencement of each 2-week randomisation block. The active follow-up versus no follow-up comparison will be evaluated as a 2×2 factorial main effect and treated as exploratory if participation rates fall below base case estimates.

### The Intervention

Women consenting to participate in the intervention will be enrolled in the B2B&Me+ mHealth coaching app and asked to schedule an appointment with a health coach through the B2B&Me+ app. Participants will be provided with Bluetooth-enabled weighing scales, Medisana BS 444 connect (26), for regular weight self-monitoring. Weight data will be transferred to the weighing scale app (which in turn is shared with the Liva Healthcare app for coaching purposes (weight data only shared if the participant provided the necessary permissions between the apps).

#### mHealth Coaching

Both synchronous and asynchronous coaching sessions will be delivered through the B2B&Me+ mHealth coaching app during the intervention (Figure 1). Two synchronous sessions will occur, the first at enrolment and second around 8 weeks postpartum. If diagnosed with GDM during pregnancy, a third session to review any health behaviour change goals to align with their diabetes management plan will be offered. Asynchronous coaching messages will be delivered according to a pre-defined schedule using a combination of both text and video. The asynchronous sessions will be weekly for the first four intervention weeks, fortnightly for two months and then monthly until birth. The postpartum sessions will be bi-weekly to support the participant to ease back into the mHealth coaching contact after birth and continue until 9 months postnatal. Session delivery will be adapted according to participant preferences.

**Fig. 1.**
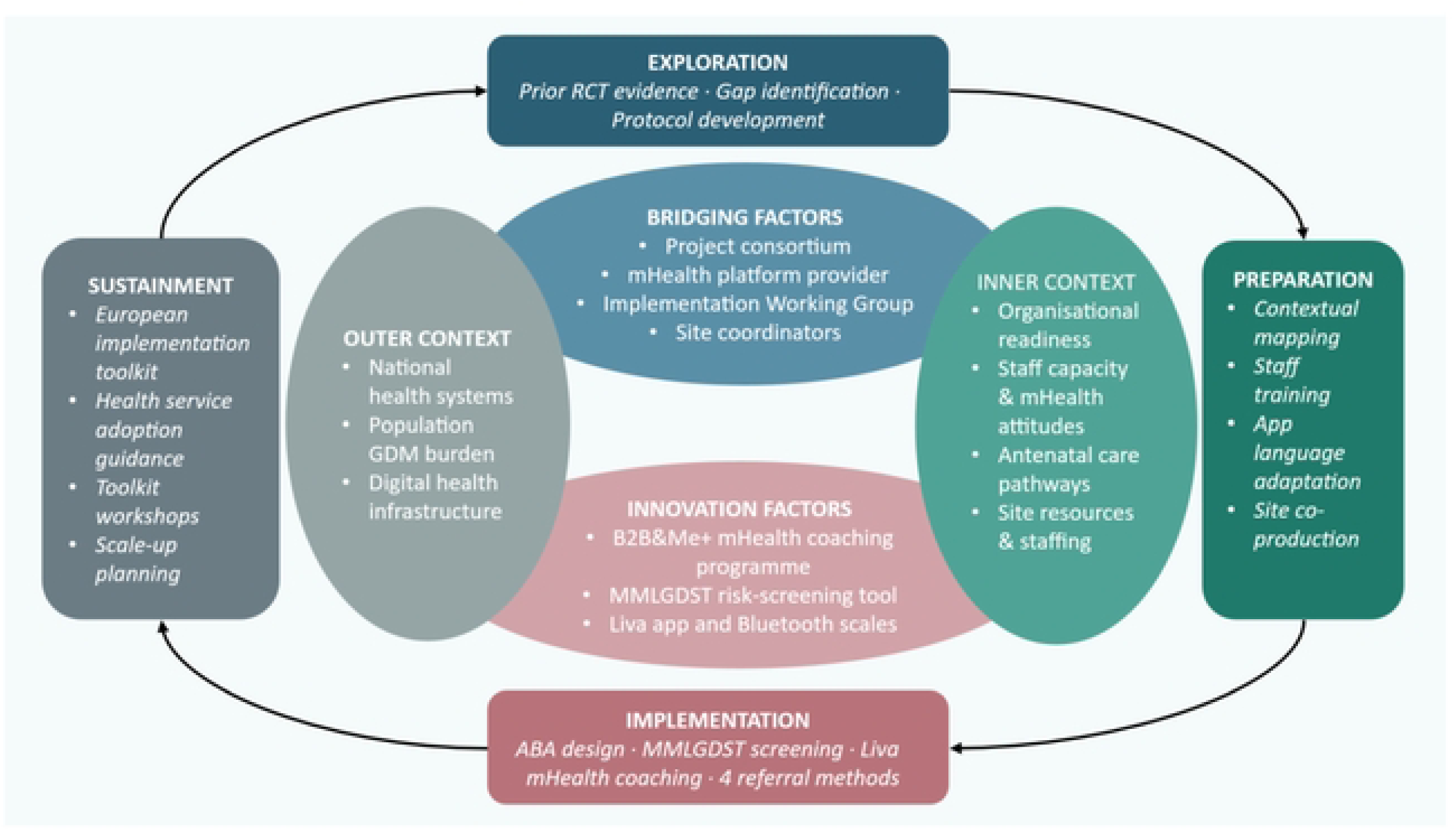
EPIS framework applied Bump2Baby and Me Plus mHealth coaching intervention (Adapted from: Aarons et al. (2011) Adm Policy Ment Health; Moullin et al. (2019) Implement Sci (19, 20))

**Fig. 2:**
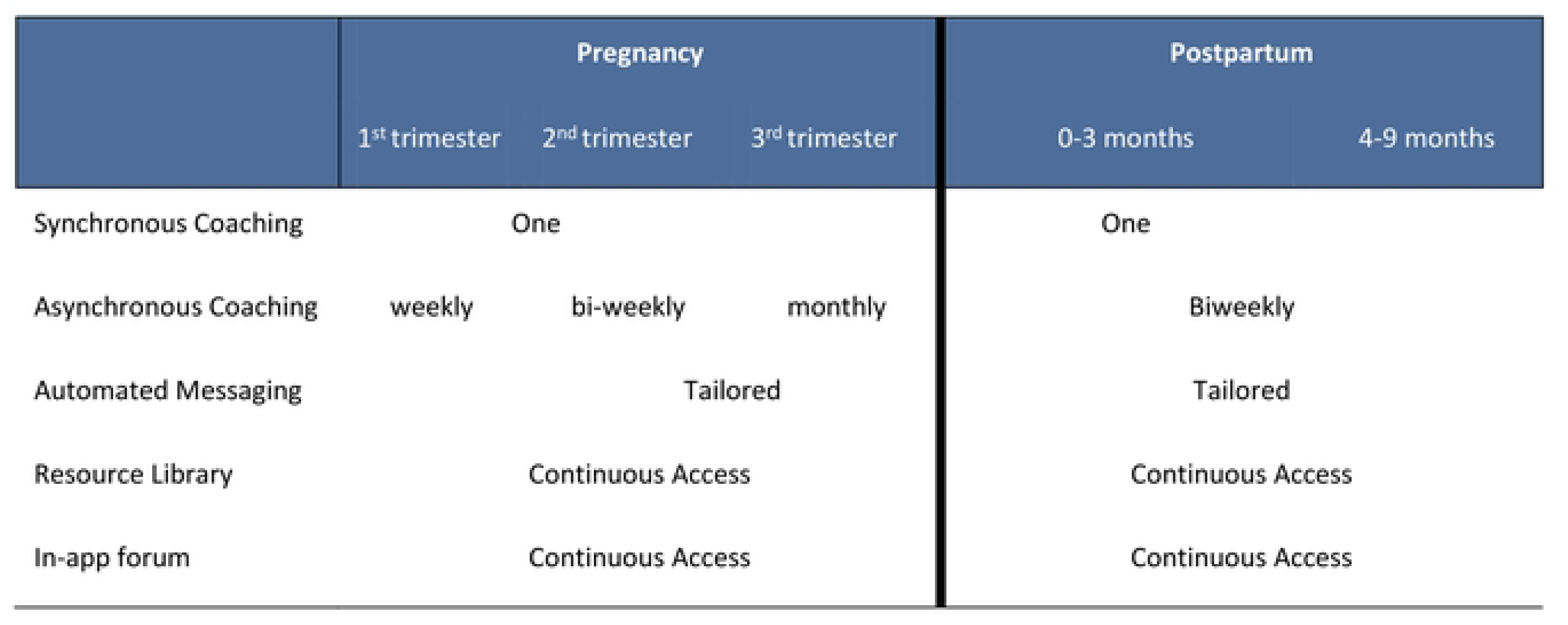
intervention Delivery.

**Fig. 3:**
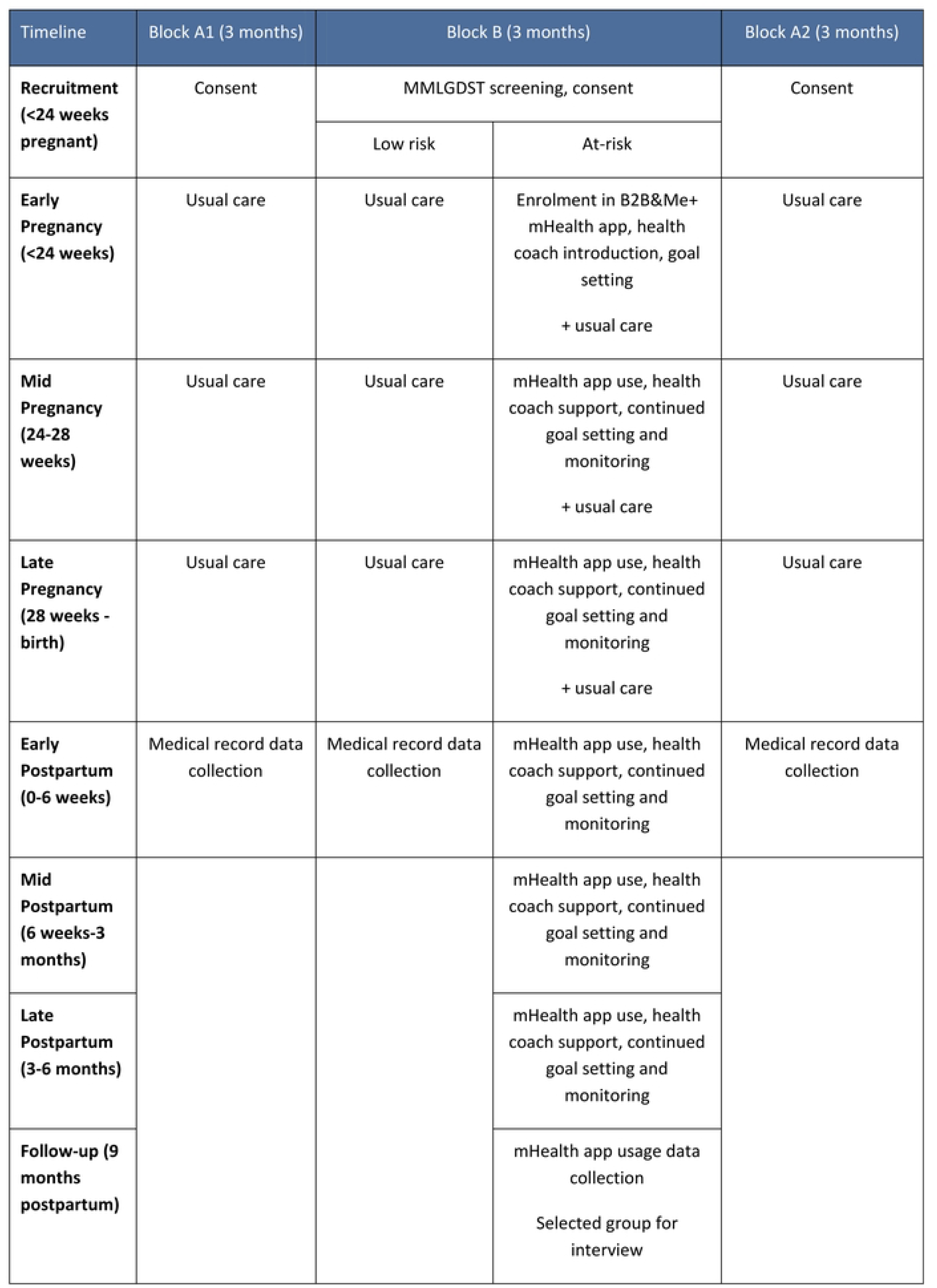
Schedule Of Evaluation Timings.

#### mHealth Resources

As part of the asynchronous messaging, participants will also receive educational resources from their health coach. This content will cover topics within healthy eating, physical activity, emotional wellbeing, breastfeeding, and best practice formula feeding. Each mHealth coach will continuously assess what content is relevant before sending it to the participants, and this content will also be freely available in the static library within the B2B&Me+ app. Follow-up messages prompting the participant to engage with the content and follow through with goals they have set for themselves will be sent through the app. The mHealth coaching platform records which of the coaching messages, educational content and goals are accessed, and this data will be used to determine the total number of messages received by message content (information, motivational, reminder). This will be used to generate an exposure index for each message type, for which the denominator will be the highest number of messages received by any one individual participant. Reminders to set goals and motivational messages once completed will be used to encourage engagement. Push notifications will be sent through the app signposting participants to additional content available to them during the weeks where no asynchronous coaching occurs. Participants will also have access to a monitored chat forum through the app to engage with other participants.

The lead development and training personnel at Liva Healthcare, alongside key study staff, will conduct the mHealth coach training. The coaching model is underpinned by a theoretical framework that draws from Social Cognitive Theory (27), Self-Determination Theory (28), and the Transtheoretical Model of Behaviour Change (29). In addition, coaches utilise Behaviour Change Technique (BCT) taxonomy, specific Motivational Intervieing Technique (MIT) taxonomy and Motivational Behaviour Change Technique (MBCT) taxonomy in order to support the participant in identifying and working towards their health and lifestyle goals. The standard Liva mHealth coach training takes a four-pronged approach, inclusive of:

1. Patient communication: including full training on the Liva coach dashboard and patient app interface.
2. Behaviour change skills: coaches are recruited with health professional backgrounds because they already have skills in supporting people to engage in behaviour change. The refresher course, and coaching handbook provided aims to build on this in line with the specific behaviour change skills used throughout Liva coaching structures.
3. Practicalities: including time management topics, remote team support and communications.
4. Patient safety and clinical pathways: medical and mental health governance structures and escalation pathways, any intervention-specific requirements.

In addition to this coaching training, all mHealth coaches will receive specific training related to the prevention and management of GDM, identification of and support for participants with postnatal depression, breastfeeding and infant feeding, infant development, and managing health for diabetes prevention.

All participants will continue to receive routine antenatal and postnatal care according to local guidelines. There are no restrictions on concomitant care or interventions, except participation in other research studies involving health behaviour change interventions during pregnancy.

At the conclusion of the study, intervention participants will receive information about continuing healthy habits and when to seek medical advice. They will also be provided with resources on local support services for maternal and child health.

### Primary Outcomes: Implementation

The implementation process will be evaluated using the RE-AIM framework (18). This framework ensures this study will collect the data necessary and adopt measures to identify the translatability and health impact of the intervention, balancing the emphasis between internal and external validity, and between individual and organisational level impact. Reach and effectiveness are individual-levels of impact, whereas adoption and implementation are organizational-levels of impact. Maintenance/sustainment can be both an individual- and an organizational-level of impact. It is pertinent to evaluate both levels because each provides valuable independent information of intervention impact (30).

Qualitative data from all interviews and focus groups will be analysed using NPT as the analytical framework to explore the mechanisms underlying RE-AIM implementation outcomes across sites.

### Secondary Outcomes: Intervention Effectiveness

Secondary outcomes will assess the effectiveness of the intervention relative to usual care only for the following outcomes:

- Change in maternal weight and BMI (gestational weight gain)
- GDM incidence
- Breastfeeding initiation and on discharge, & breastfeeding duration
- Diet quality
- Physical activity
- Rates of adverse birth outcomes (pre-eclampsia, preterm birth, birth weight ≥4000g, birth trauma, neonatal respiratory distress, phototherapy, stillbirth/neonatal death, NICU admission or shoulder dystocia, caesarean section/interventions)
- Healthcare utilisation

### Data on Participants

Data will be collected from all participants using a questionnaire administered at baseline recruitment. Medical records will be used to validate this information and to track the participants throughout the pregnancy, birth, and up to postnatal discharge. Data will also be sourced from the mHealth app for those within the intervention, which will include health and behaviour outcomes, and statistics on their engagement with the app and the health coach (Table 2).

**Table 1:**
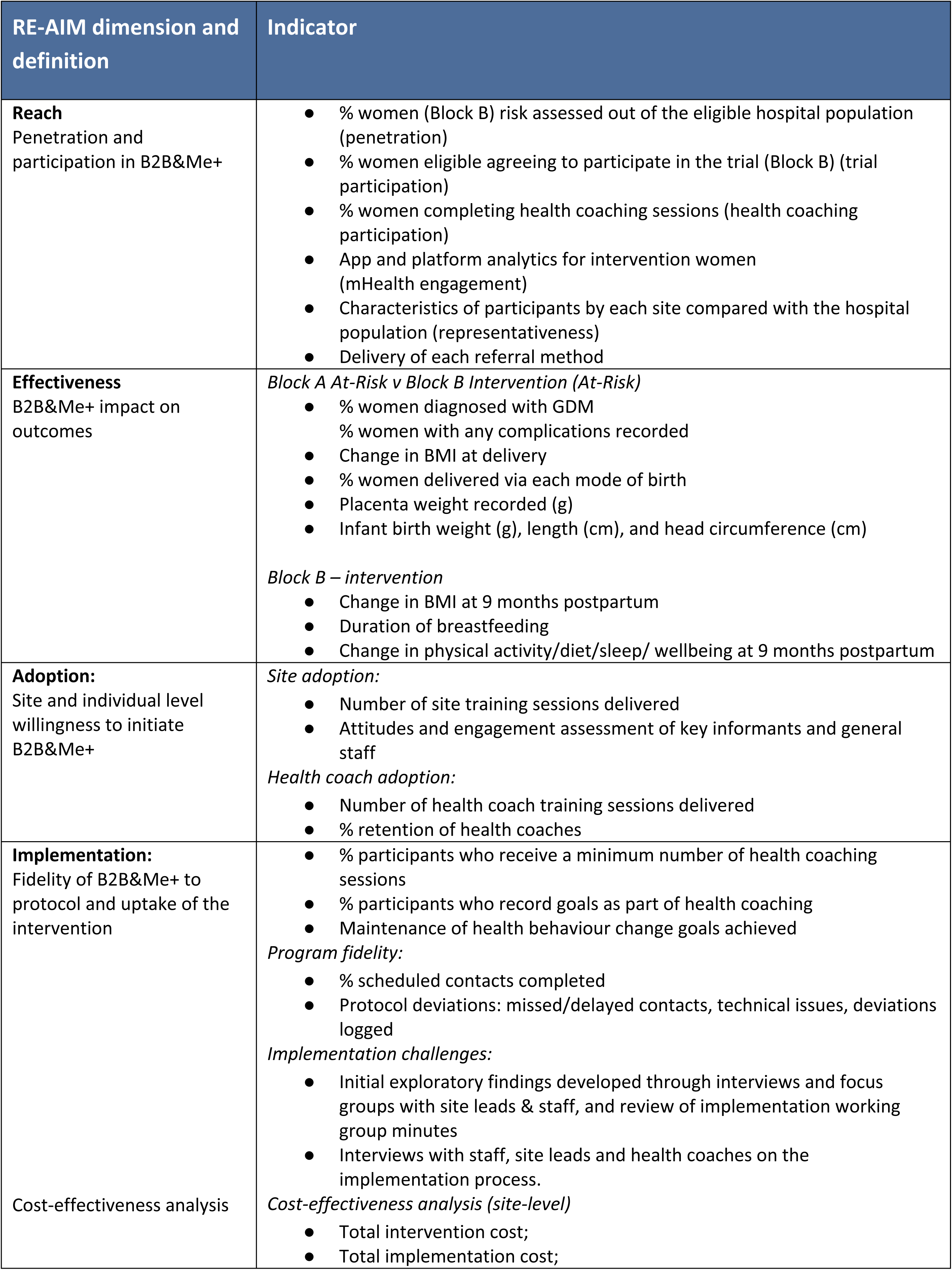
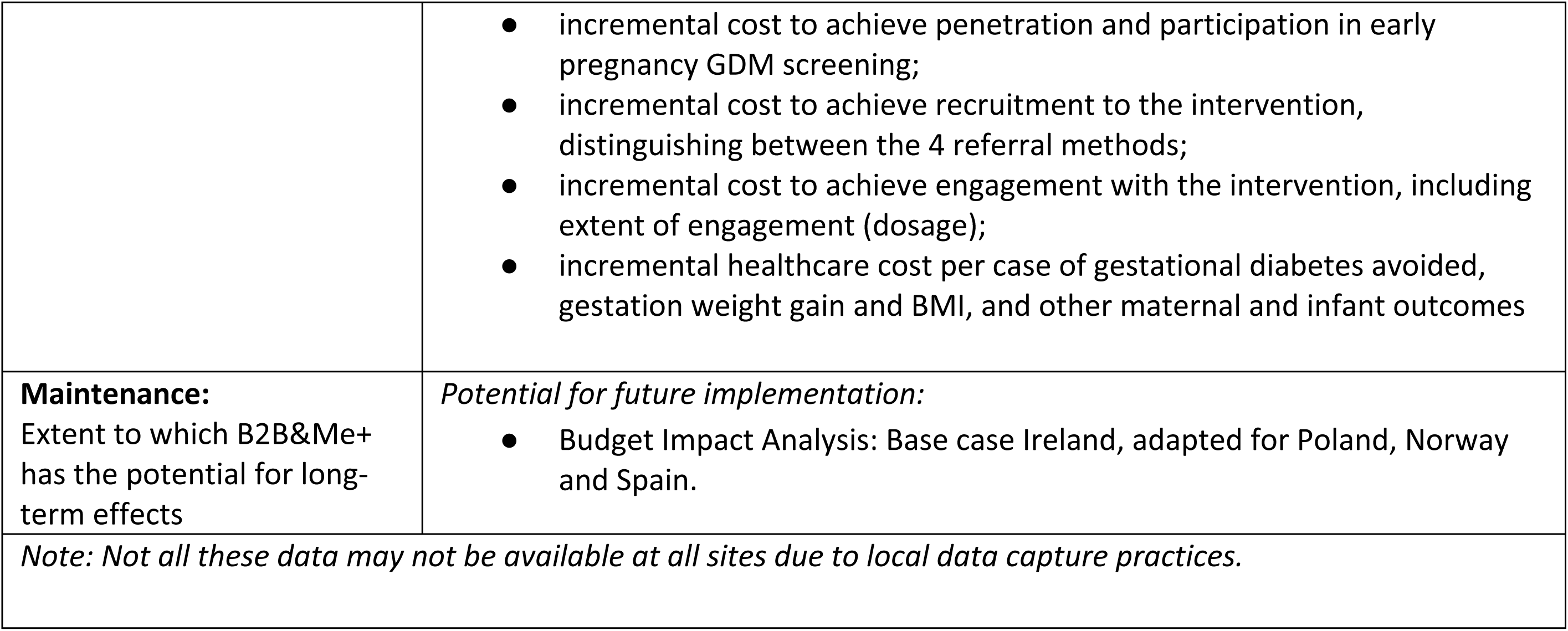
RE-AIM dimensions and indicators.

| RE-AIM dimension and definition | Indicator |
| --- | --- |
| **Reach**<br>Penetration and participation in B2B&Me+ | • % women (Block B) risk assessed out of the eligible hospital population (penetration)<br>• % women eligible agreeing to participate in the trial (Block B) (trial participation)<br>• % women completing health coaching sessions (health coaching participation)<br>• App and platform analytics for intervention women (mHealth engagement)<br>• Characteristics of participants by each site compared with the hospital population (representativeness)<br>• Delivery of each referral method |
| **Effectiveness**<br>B2B&Me+ impact on outcomes | *Block A At-Risk v Block B Intervention (At-Risk)*<br>• % women diagnosed with GDM<br>• % women with any complications recorded<br>• Change in BMI at delivery<br>• % women delivered via each mode of birth<br>• Placenta weight recorded (g)<br>• Infant birth weight (g), length (cm), and head circumference (cm)<br>*Block B – intervention*<br>• Change in BMI at 9 months postpartum<br>• Duration of breastfeeding<br>• Change in physical activity/diet/sleep/ wellbeing at 9 months postpartum |
| **Adoption:**<br>Site and individual level willingness to initiate B2B&Me+ | *Site adoption:*<br>• Number of site training sessions delivered<br>• Attitudes and engagement assessment of key informants and general staff<br>*Health coach adoption:*<br>• Number of health coach training sessions delivered<br>• % retention of health coaches |
| **Implementation:**<br>Fidelity of B2B&Me+ to protocol and uptake of the intervention<br>Cost-effectiveness analysis | • % participants who receive a minimum number of health coaching sessions<br>• % participants who record goals as part of health coaching<br>• Maintenance of health behaviour change goals achieved<br>*Program fidelity:*<br>• % scheduled contacts completed<br>• Protocol deviations: missed/delayed contacts, technical issues, deviations logged<br>*Implementation challenges:*<br>• Initial exploratory findings developed through interviews and focus groups with site leads & staff, and review of implementation working group minutes<br>• Interviews with staff, site leads and health coaches on the implementation process.<br>*Cost-effectiveness analysis (site-level)*<br>• Total intervention cost;<br>• Total implementation cost; |
|  | • incremental cost to achieve penetration and participation in early pregnancy GDM screening;<br>• incremental cost to achieve recruitment to the intervention, distinguishing between the 4 referral methods;<br>• incremental cost to achieve engagement with the intervention, including extent of engagement (dosage);<br>• incremental healthcare cost per case of gestational diabetes avoided, gestation weight gain and BMI, and other maternal and infant outcomes |
| **Maintenance:**<br>Extent to which B2B&Me+ has the potential for long-term effects | *Potential for future implementation:*<br>• Budget Impact Analysis: Base case Ireland, adapted for Poland, Norway and Spain. |
| *Note: Not all these data may not be available at all sites due to local data capture practices.* |  |

**Table 2:** Data collected from participants.

| Data Collected from Participants |  |
| --- | --- |
| ALL PARTICIPANTS |  |
| Data collected at Baseline<br>(Questionnaire & cross-check against medical records) |  |
| Demographic characteristics | Clinical information |
| • Age<br>• Ethnicity<br>• Educational attainment<br>• Relationship status | • Gestational age at inclusion<br>• Relevant medical history including PCOS diagnosis<br>• Obstetric history (gravidity, parity, previous complications)<br>• Prior GDM screening and results<br>• Anthropometric measurements (weight and height)<br>• Blood pressure |
| Data extracted from Medical Records |  |
| Antenatal Assessments | Birth and postnatal |
| • 20-week scan details<br>• Glucose tolerance testing (<24 weeks & 24-28 weeks)<br>• Gestational diabetes diagnosis<br>• Antenatal visit record<br>• Admissions (unplanned/planned)<br>• GDM treatment type | • Gestational age at delivery (weeks)<br>• Weight of woman (collected closest to delivery)<br>• Delivery method, including any medical interventions<br>• Birth complications, including admission to the NICU, special care nursery and/or neonatal hypoglycaemia<br>• Placental weight<br>• Apgar score<br>• Newborn weight<br>• Newborn length<br>• Newborn head circumference<br>• Breastfeeding initiation<br>• Breastfeeding status on discharge<br>• Lactation support services accessed<br>• Blood results for anaemia investigations and glycaemia status<br>• Healthcare utilisation (appointments, admissions – as captured on hospital records) |
| Participants in Intervention Only |  |

| Assessments recorded in the B2B&Me+ app |  |  |
| --- | --- | --- |
| At recruitment | Periodically throughout the intervention<br>(as uploaded by participant) | 9 months postpartum |
| FIGO nutrition checklist scores<br>WHO-5<br>MMLGDST classification | Weight (recorded using the Bluetooth scales provided)<br>Steps/Activity tracking | FIGO nutrition checklist scores<br>WHO-5 |
| Adherence to the intervention will be monitored through app usage metrics, completion of self-monitoring activities, and engagement with the health coach. |  |  |
| *Note: Not all these data may not be available at all sites due to local data capture practices.* |  |  |

**Table 3:**
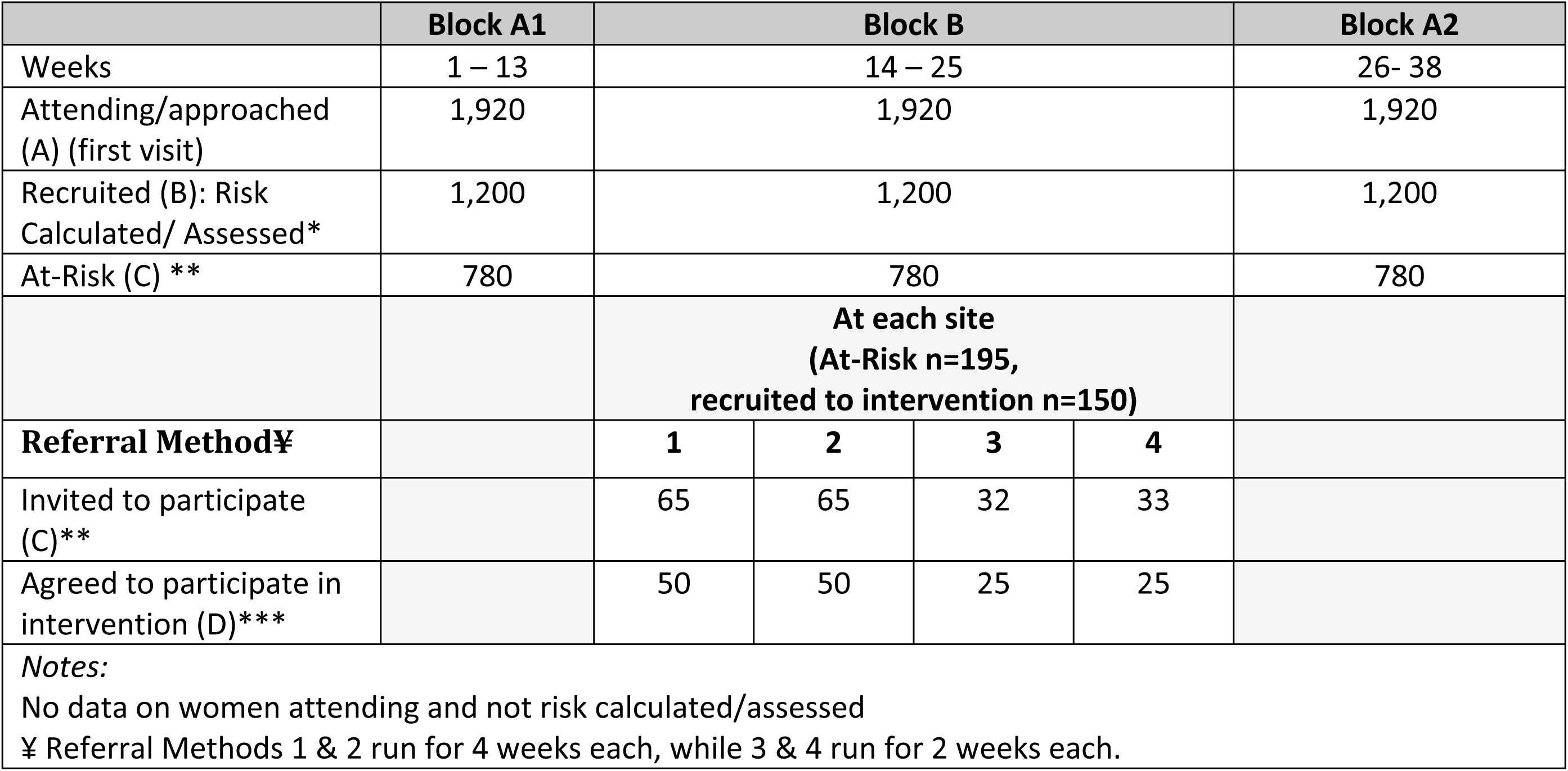
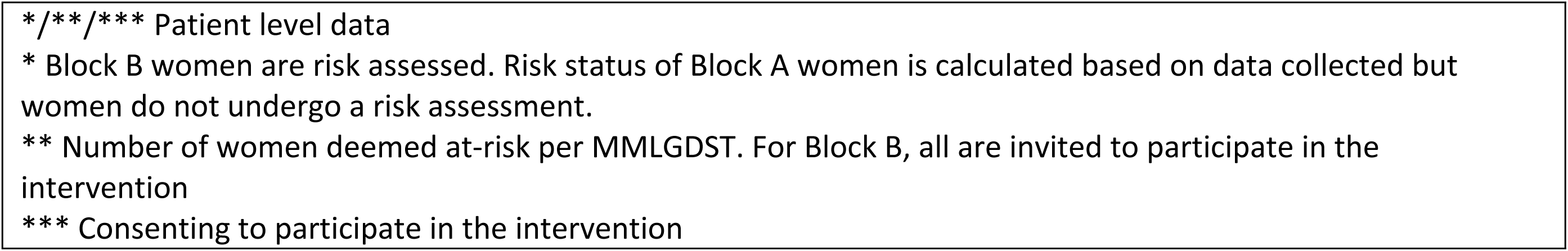
Expected participant recruitment.

|  | Block A1 | Block B |  |  |  | Block A2 |
| --- | --- | --- | --- | --- | --- | --- |
| Weeks | 1 – 13 | 14 – 25 |  |  |  | 26- 38 |
| Attending/approached (A) (first visit) | 1,920 | 1,920 |  |  |  | 1,920 |
| Recruited (B): Risk Calculated/ Assessed* | 1,200 | 1,200 |  |  |  | 1,200 |
| At-Risk (C) ** | 780 | 780 |  |  |  | 780 |
|  |  | At each site<br>(At-Risk n=195,<br>recruited to intervention n=150) |  |  |  |  |
| **Referral Method¥** |  | **1** | **2** | **3** | **4** |  |
| Invited to participate (C)** |  | 65 | 65 | 32 | 33 |  |
| Agreed to participate in intervention (D)*** |  | 50 | 50 | 25 | 25 |  |
| **Notes:**<br>No data on women attending and not risk calculated/assessed<br>¥ Referral Methods 1 & 2 run for 4 weeks each, while 3 & 4 run for 2 weeks each. |  |  |  |  |  |  |

\*/\*\*/\*\* Patient level data \* Block B women are risk assessed. Risk status of Block A women is calculated based on data collected but women do not undergo a risk assessment. \*\* Number of women deemed at-risk per MMLGDST. For Block B, all are invited to participate in the intervention \*\*\* Consenting to participate in the intervention

Data collected will be entered into an online, secure platform with data stored on a secure server located at the lead institution. The data entry platform will be accessible to research staff through the use of individual log-ins. Local researchers will have access restricted to their own site. Data captured in the database will be pseudonymised, with all patients assigned a Study ID.

### Economic evaluation

As a hybrid type 3 study, the cost effectiveness of both running and implementing the intervention will be assessed for sites individually and collectively as follows:

i. The incremental cost of the intervention compared to usual care per health outcome achieved (see above)
ii. The incremental total cost (intervention + implementation) per health outcome achieved.

#### Intervention cost-effectiveness analysis

The initial cost-effectiveness analysis (CEA) will calculate gains in treatment effect and costs of the intervention and compares the treatment effects and costs to the comparator, usual care. The product of the cost-effectiveness analysis is an incremental cost-effectiveness ratio (ICER) which expresses the additional costs per additional outcome gained for the intervention compared to usual care for pregnant women at risk of gestational diabetes.

#### Intervention Implementation CEA

The capture of costs must be sufficiently comprehensive to ensure the cost of fully replicating the intervention is measured. As the treatment and reporting of costs and outcomes in this project must facilitate implementation to other settings – scaling up (to similar organizations or systems) and scaling out (to different types of settings and locations), reporting will focus on replicability through full transparency in results (31). Implementation costs include intervention replication costs that would be required by those adopting the program (32), costs related to intervention tailoring or adaptation, costs to engage practitioners and participants, and any other costs related to implementation strategies such as training, supplies and labour costs for activities such as provider coaching or consultation. Costs and outcomes will be reported at site-level, with the local contextual factors likely to influence results identified. Intervention costs (e.g., coaching support, app costs, etc.) will be captured, as will implementation costs, many of which are activity-driven based on time taken to complete implementation tasks (33). Research-specific activities that are essential to designing and developing the original implementation will be excluded (31, 34). Participant and staff perspectives on the acceptability, feasibility and usefulness of the intervention and its fidelity will be gathered using focus groups and/or interviews in order to better understand the full story of the costs and outcomes of implementation. Results from the economic evaluation will inform more widespread scale-up of this intervention.

#### Referral CEA

Comparative economic evaluation of implementation strategies provides critical information for payers, policymakers, and providers to make informed decisions if specific strategies are an efficient use of scarce organizational resources (35). A systematic evaluation of each implementation strategy and the costs of achieving them will be undertaken (33). Cost-effectiveness analysis will assess variation in recruitment between each of the four referral methods. These referral methods vary in their resource requirements: leaflet versus point of care (POC) referral, and both options accompanied by a follow-up phone call. Capturing implementation costs will involve the completion of a tracking form (Supplementary File 2: Table A4) by each site that will record the time spent and costs incurred recruiting women to the intervention, distinguishing between research costs (e.g. obtaining consent), intervention and implementation costs common to all four recruitment methods (e.g. completing the risk assessment), and time and costs specific to each of the four recruitment strategies. To minimise the burden of data collection, the template will be completed for a sample of five consecutive participants recruited to the intervention per 2-week block. Site-specific pay scales for staff involved in the delivery and/or implementation of the intervention in a real-world context will be used to estimate activity-based estimates of the cost of conducting each specified step of the four recruitment strategies. The incremental cost of implementing each of the four recruitment strategies, with *Leaflet Only* as the base case, will then be assessed against the relative recruitment success.

#### Budget Impact Analysis

A budget impact analysis (BIA) will estimate changes in the expenditure incurred by the healthcare system after adoption of B2B&Me+. This approach suits the implementation science context where financial factors are central to whether programs and services are adopted and sustained (35). Key questions include how effective is the implementation of the intervention in reaching the target population, the staffing and resources required to meet the new demand for services, and the budget required going forward (31). The BIA takes a broad perspective to include the entire healthcare organisation as beneficiaries and assess the downstream budget impacts: costs saved via preventive care (36). The Irish Healthcare system will be the initial base case for the BIA.

### Sample Size

Observed data from Block A1 across all four sites confirm that 65% of women screened using the MMLGDST are classified as at-risk for GDM, yielding approximately 195 eligible women per block per site. Of these 20% are anticipated to decline intervention participation, which is roughly half the number of women declining participation for the B2B&Me RCT where randomisation to control arm existed. On this basis, we expect to recruit approximately 150 intervention participants per site and 600 across all sites. The at-risk women identified during the two usual care blocks (Blocks A1 and A2) constitute the comparison group, yielding n=1,560 comparators (195 at-risk × 2 blocks × 4 sites). For the primary clinical effectiveness outcome (GDM incidence), we powered based on observed rates from the B2B&Me RCT, where GDM was diagnosed in 14.0% of intervention participants (53/379) compared with 19.7% of control (75/380; p=0.034)(15). With n=600 intervention and n=1,560 comparison group, the study will have 88.4% power to detect a difference of this magnitude at a two-sided significance level of 0.05, with an acceptable false negative rate of 0.116 (37).

The referral method comparison is structured as a 2×2 factorial design, with referral mode (point-of-care versus information leaflet) and active follow-up (follow-up phone call versus no follow-up call) as the two independent factors. Anticipated participation rates are derived from Porter et al. (2021), who reported pooled participation rates of 46.6% for point-of-care approaches and 11.5% for leaflet approaches across a comparable hybrid type 3 referral methods study in primary care (21). For the primary referral method comparison (point-of-care versus leaflet), applying a conservative 50% attenuation of these rates to account for population and setting differences, and a design effect of 1.5 to account for clustering by site, a minimum of 95 women per group is required to detect this difference with 80% power at α=0.05. With approximately 520 women anticipated in the point-of-care group and 256 in the leaflet group across four sites, this comparison is robustly powered under all scenarios tested. For the secondary referral method comparison (active follow-up versus no follow-up), Porter et al. observed pooled participation rates of 22.5% and 12.5% respectively. Applying a design effect of 1.5, a minimum of 338 women per group is required at base case rates, with approximately 388 available in each group. This comparison is adequately powered under base case assumptions; however, if participation rates fall below those observed by Porter et al., findings from this comparison will be treated as exploratory. Results will be reported with 95% confidence intervals throughout to support interpretation irrespective of formal significance.

### Statistical Analysis

#### Primary Analysis

Penetration and participation are important measures for the assessment of implementation. Participation is calculated as the number of participants recruited to the intervention (D) expressed as a proportion of those eligible and invited (C), while penetration is calculated based on the number of participants screened (B) expressed as a percentage of those attending the clinic and eligible for screening (A). The overall difference in penetration between the intervention and control groups and difference in participation rates across the four referral methods will be assessed. While penetration is not directly related to referral method, there may be a spillover staffing effect due to the level of resource taken to recruit to the intervention being higher for POC than leaflet recruitment, resulting in less time available for staff to conduct assessments. Therefore, penetration will also be assessed by referral method. The level of engagement by intervention participants will also be assessed, as defined by the extent of engagement with the health coach and the app. Chi-square tests will be used for categorical data and t-tests or ANOVA for continuous data.

#### Secondary analysis

Secondary analysis will compare the effectiveness of the intervention across different blocks (A1 & A2 (at-risk) vs B (intervention)) and by site. This will be analysed using chi-square tests for categorical data and t-tests or ANOVA for continuous data. Non-normally distributed data will be analysed either after log transformation or with non-parametric methods. Linear regression analysis will account for potential confounding variables. Dichotomous outcomes will be analysed using logistic regression. Outcomes with multiple measurements at pre-defined time points will be analysed using mixed models with generalised estimating equations (GEE), with either a linear or a logistic link function, depending on the nature of the outcome.

Missing data will be handled using multiple imputation techniques where appropriate, with sensitivity analyses used to assess the impact of missing data assumptions. Pattern-mixture models will be used to explore potential non-random missing data mechanisms (38). Sensitivity analyses will also be conducted to assess the robustness of findings to different analytical approaches and assumptions.

#### Subgroup analyses

Subgroup analyses will be conducted to examine potential effect modifiers, including maternal age, pre-pregnancy BMI categories, parity (nulliparous vs multiparous), socioeconomic status, site/country differences, referral method (within Block B).

### Implementation Toolkit

On conclusion of the analysis, a European implementation toolkit will be developed to provide health services with scalable strategies to bridge the evidence-to-practice gap. The toolkit will be co-designed with all key stakeholders and aims to support healthcare services and systems looking to implement and scale up the B2B&Me+ intervention. It is planned to contain a road map on stakeholder engagement across management and frontline staff, project governance recommendations, training resources and implementation and delivery framework, recommendations and resources targeting staff, women and managers (including resources required and anticipated savings). Measurement of fidelity, adaptability and evaluation will also be covered, as will strategies to link to audit and feedback with routine clinical data sets for ongoing evaluation and improvement.

## DISCUSSION

This protocol outlines a multi-country, hybrid type 3 implementation effectiveness study evaluating the integration of the B2B&Me+ programme into routine maternity care. This study will address a key gap in understanding how to translate evidence from clinical trials into routine practice and sustain interventions in real-world healthcare systems. Implementation outcomes will be prioritised while concurrently assessing effectiveness within diverse clinical contexts. The hybrid type 3 design brings a primary emphasis on implementation outcomes, while examining effectiveness in relation to intervention uptake and fidelity. This pragmatic approach enhances both internal and external validity and policy relevance.

The study is further strengthened by its theoretical grounding in the RE-AIM framework, which will guide the evaluation of reach, effectiveness, adoption, implementation and maintenance. In particular, the inclusion of adoption and maintenance outcomes will provide important insights into the sustainability and scalability of the intervention beyond the study period. The integration of qualitative methods and quantitative measures will allow for exploration of contextual determinants, supporting a more nuanced understanding of how and why implementation succeeds or falls short across settings.

The multi-country design strengthens the study by enabling the examination of implementation across heterogeneous maternity healthcare systems, organisational structures, and cultural contexts. Such cross-contextual evaluation is critical for obtaining and advancing generalisable knowledge in implementation science. By incorporating contextual mapping and stakeholder engagement, the study will identify key barriers and facilitators at multiple levels, including participant, provider and system levels. These findings will inform and benefit the design of a European implementation toolkit, which has the potential to support the transferability and adaptation of the intervention across different settings.

The evaluation of different referral strategies provides an additional level of innovation to optimise intervention reach and engagement. By systematically comparing point-of-care and more passive referral approaches, with and without follow-up contact, the study will generate evidence on mechanisms influencing participation. Understanding how referral processes affect uptake is critical, particularly for preventive interventions, where engagement may often be considered suboptimal.

Despite these strengths, several limitations should be acknowledged. The non-randomised ABA design introduces the potential for temporal confounding and limits causal inference compared to fully randomised designs. Variability in usual care across sites may also influence observed differences in outcomes. Although analytical strategies will seek to account for these factors, residual confounding cannot be excluded. Additionally, relying on a digital health platform may introduce selection bias, as participation requires engagement with smartphone technology, potentially excluding more vulnerable populations. Limiting inclusion to women speaking the country’s language also excludes more vulnerable populations. We would hope to include these populations should the use of the app be further expanded.

Implementation challenges are also anticipated, including variability in organisational readiness, competing clinical demands and differences in infrastructure across sites. Achieving an appropriate balance between intervention fidelity and context-specific adaptation will be essential, particularly in a multi-country context. While training, monitoring and process evaluation are incorporated to mitigate these challenges, variation in implementation is likely and should be viewed as both a limitation and an important, active source of learning. Furthermore, sustaining participant engagement with mHealth coaching across the perinatal period may prove challenging and entail implications for both implementation and effectiveness outcomes.

The findings from this study will have significant relevance and implications for both practice and policy. In providing evidence on the feasibility, acceptability, and cost-effectiveness of integrating a digitally supported health behaviour intervention into routine maternity care, the study will inform decision-making regarding scalability and sustainability. The development of an implementation toolkit, based on data captured across multiple European contexts, will provide practical guidance for healthcare systems seeking to incorporate interventions for GDM and related conditions.

In conclusion, this study will contribute to the advancement of implementation science by providing rigorous, informed evidence on the processes and outcomes associated with integrating an evidence-based intervention into routine maternity care. Through its hybrid design, multi-level evaluation, and cross-context scope, the B2B&Me+ project is well-positioned to generate transferable insights that can inform the implementation of interventions in maternal health and beyond. Recruitment has already commenced, with Block A1 beginning in December 2025, and overall (Blocks A2 & B) recruitment scheduled to conclude in August 2026.

### Ethical Approval

Primary research ethics approval has been obtained from the National Maternity Hospital and University College Dublin in Dublin, Ireland (15^th^ April 2025). Site specific study approval has also been obtained from each study site’s relevant human research ethics boards as follows:

- Granada: Bioethical Committee for Clinical Research of San Cecilio University Clinical and Mother-Infant Hospitals of Granada (Spain) (5^th^ October 2025).
- Poland: Bioethics Committee at the Pomeranian Medical University in Szczecin (25^th^ June 2025).
- Norway: Regional Committee for Medical and Health Research Ethics South-East Norway (16th October 2025).

### Patient and Public Involvement

Women with a recent history of obesity/overweight or GDM and pregnancy will be invited to participate as experts by lived experience advisory panel. This panel will form the Bump2Baby and Me+ patient and public involvement (PPI) group. The PPI panel will comprise 3-4 representatives from each site who will attend an online meeting twice a year conducted in English that will last a maximum of 1 hour. Members are expected to remain on the panel for the duration of the project. The panel can advise on all participant-facing aspects of the project, with the aim of ensuring the voice of the woman remains at the centre of the project. UCD will maintain and provide administrative support to the PPI panel.

## Data Availability

No datasets were generated or analysed during the current study. All relevant data from this study will be made available upon study completion.

## Funding

This project has received funding from the Health Research Board, Innovation Fund Denmark, Norges Forskningsråd, Narodowe Centrum Badań i Rozwoju, and Consejería de Salud y Consumo de la Junta de Andalucía under the framework of the co-fund partnership Transforming Health and Care Systems (THCS, GA N° 101095654) of the EU Horizon Europe Research and Innovation Programme. **Disclaimer.** The opinions and views expressed represent those of the author and do not necessarily represent those of the funders, who cannot be held liable for them.

## Abbreviations

GDM: Gestational diabetes mellitus
MMLGDST: Monash machine learning GDM screening tool
T2D: Type 2 diabetes
BMI: Body mass index
GWG: Gestational weight gain
B2B&Me: Bump to Baby and Me
POC: Point of Care
mHealth: Mobile health
EPIS Framework: Exploration, Preparation, Implementation, Sustainment Framework
NPT: Normalisation Process Theory
RE-AIM framework: Reach, Effectiveness, Adoption, Implementation, Maintenance framework
CEA: Cost-effectiveness analysis
ICER: Incremental cost-effectiveness ratio
BIA: Budget impact analysis
ANOVA: Analysis of Variance
GEE: Generalised estimating equations
PPI: Patient and public involvement
WHO: World Health Organisation
FIGO: International Federation of Gynaecology and Obstetrics
PCOS: Polycystic Ovary Syndrome

## Notes

### Competing Interest Statement

The authors have declared no competing interest.

### Clinical Trial

NCT07189221

### Author Declarations

Primary research ethics approval has been obtained from the National Maternity Hospital and University College Dublin in Dublin, Ireland (15th April 2025). Site specific study approval has also been obtained from each study site’s relevant human research ethics boards as follows: ? Granada: Bioethical Committee for Clinical Research of San Cecilio University Clinical and Mother-Infant Hospitals of Granada (Spain) (5th October 2025). ? Poland: Bioethics Committee at the Pomeranian Medical University in Szczecin (25th June 2025). ? Norway: Regional Committee for Medical and Health Research Ethics South-East Norway (16th October 2025).

